# Efficacy of Electronic Travel Aids for the Blind and Visually Impaired During Wayfinding

**DOI:** 10.1101/2025.07.07.25330998

**Authors:** Claire E. Pittet, Eduardo Villar Ortega, Maël Fabien, Mark T. Wallace, Monica Gori, Micah M. Murray

## Abstract

Independent traveling remains challenging for blind and visually impaired (BVI) individuals. While the white cane is effective at detecting ground-level obstacles, it provides no information about elevated obstacles or object characteristics. Recent technologies have been designed to support navigation as well as object detection. In our study, we compared the performance of 13 BVI participants who separately used two secondary electronic travel aids (ETAs) versus cane use alone. One ETA was a camera-based mobility vest (NOA), and the other was an ultrasonic sensor-based wearable (BuzzClip). Participants completed an obstacle avoidance task with both ETAs and an object detection task using two versions of NOA’s object-finding functionality. Quantitative performance measures and semi-structured interviews were collected. NOA resulted in enhanced obstacle avoidance. Participants used their canes less and collided less with obstacles when using NOA than the BuzzClip or the white cane alone. NOA resulted in lower frustration and higher perceived performance, as well as greater perceived safety and obstacle detection than the BuzzClip. Object-finding performance outcomes were similar across both versions, suggesting potential benefit from a dynamic combination of approaches tailored for each user. Collectively, these data underscore how ETAs may be integrated into use by the BVI community.

## 1 Introduction

Blind and visually impaired (BVI) individuals must overcome multiple obstacles when navigating urban environments. Unpredictable static and moving obstacles, crowded areas, and complex pathways significantly limit their ability to move in an independent manner [1–3]. The inability to navigate independently has serious consequences for both the autonomy and safety of BVI individuals. For example, in a survey of over 300 BVI individuals, 13% stated they experienced head injuries monthly or more frequently and 7% reported falling at least once a month due to such hazards [4].

Given these risks, mobility aids are devices that support independent navigation for blind and visually impaired individuals, ensuring both safety and independence. They include both primary aids, such as the white cane and guide dog, and secondary aids, which serve as supplementary tools to enhance primary aids. Many secondary aids are electronic travel aids (ETAs) that use sensors and feedback—typically ultrasonic, infrared, or camera-based—to detect obstacles and convey spatial information through auditory or haptic cues. While ETAs are most often used alongside primary aids, certain devices, such as electronic canes, can also function as primary aids.

Currently, the white cane remains the most widely used tool. It is an affordable, recognizable, and accessible solution for many people. However, the white cane also has certain limitations. It is limited in its coverage to the length of the cane itself and to the surface(s) on which it is being swiped (typically the ground). As such, the white cane does not protect users from obstacles at the upper body and head levels, which can sometimes result in hazardous collisions [4]. Furthermore, even with optimal swiping techniques, the white cane rarely provides full mobility coverage [5,6]. While the white can convey tactile cues about the contact surface (e.g., texture, hardness), it does not provide explicit semantic information about what objects are, nor about objects beyond its physical reach. Nor does the white cane provide navigational guidance; the user needs to know their route. It also provides no information about objects approaching unless the user is swiping, which can result in collisions when they are not.

Examples of ETAs include smart canes such as the WeWALK (WeWALK, https://www.wewalk.io) and the Ultracane (Sound Foresight Technology, https://www.ultracane.com) integrate ultrasonic sensors directly into a cane and provide haptic or audible feedback. Clip-on devices such as Rango (GoSense, https://www.gosense.com) and the BuzzClip (previously developed by iMerciv, currently no longer available) attach to a white cane and provide obstacle alerts via spatialized audio or vibrations. Handheld devices like the MiniGuide (GDP Research, https://www.gdp-research.com.au) also use ultrasonic sensors and provide haptic cues, while wearables such as the Sunu Band (previously developed by Sunu Inc., now discontinued) provide haptic feedback to the wrist.

More recently, advanced systems such as NOA (biped robotics, https://www.biped.ai), Glide (Glidance Inc., https://glidance.io), or .lumen Glasses (.lumen, https://www.dotlumen.com) have emerged. These newer systems use computer vision and artificial intelligence (AI) to enhance obstacle detection, which allows them to go even further than ultrasonic systems. As of today, only NOA is commercially available; Glide and the .lumen glasses remain at the prototype stage. Permanent Internet Archive links are provided in Table S1 to ensure long-term accessibility, even if the original websites become unavailable.

While promising, the effectiveness of ETAs remains inconsistent and has not been empirically validated. Many limitations stem from technological constraints, challenges in user experience, and practical application constraints; all of which must be examined rigorously to ensure user acceptance [7–9]. According to [7], out of the 140 studies on ETAs they reviewed, only 29**\**% included BVI participants. This low proportion significantly undermines the validity of such evaluations. In many cases, tests are limited to prototype-level experiments of technical functionality without involving end-users at all [10–12]. Moreover, when testing does occur, it often relies on sighted blindfolded individuals, who lack the orientation and mobility skills from BVI individuals [13–15]. These approaches do not yield assessments that reflect real-world usability [16]. It is therefore crucial for current and future research to be conducted in a user-centered manner by directly involving BVI participants.

In this context, the present study quantitatively and qualitatively evaluates two specific secondary ETAs: the BuzzClip and NOA devices (Fig. 1). The BuzzClip was chosen as a representative example of common ultrasonic-based ETAs. NOA was selected as a novel device of interest, whose performance we aimed to assess. Both devices aim to help BVI individuals navigate urban environments safely. The BuzzClip uses ultrasound and haptic feedback to signal hazards at various heights. NOA, equipped with three cameras and AI, aims to do the same by providing spatialized auditory feedback about obstacles. Here, we compare the BuzzClip and NOA devices to determine how each impacts obstacle detection, mobility performance, and perceived workload when used alongside a white cane. This design allows us to evaluate the potential added value of each ETA by comparing their combined use with a white cane against the baseline condition of using the cane alone, reflecting the participants’ usual mobility practices.

**Figure 1.**
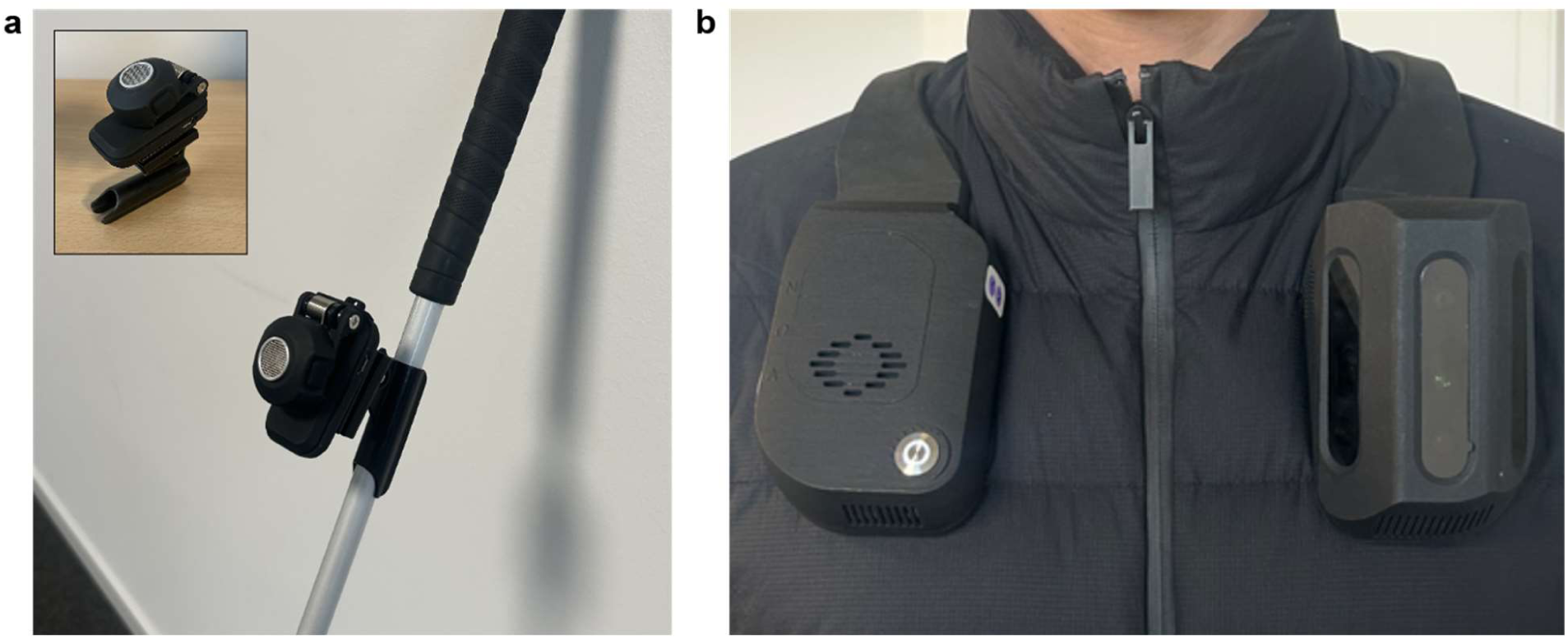
Images of the two ETAs evaluated in the study. (**a**) The BuzzClip emits vibrations and is attached to a white cane. (**b**) The NOA device provides spatialized audio guidance and is worn on the shoulders.

Additionally, in a second task, a new object-finding feature on the NOA device is tested with BVI participants. This novel feature is independent of the obstacle detection functionality tested in the first task and focuses on locating and guiding users to a target object or landmark. To better understand users’ needs and optimize system design, two versions are compared: one based on generative AI and the other on deep learning (DL). While the generative AI version provides more natural, descriptive language, the DL version focuses on precise, actionable spatial guidance.

For both tasks in the study, we collect both qualitative feedback from participants and quantitative performance metrics. Quantitative performance metrics include walking speed (used to derive the preferred percentage of walking speed), cane contacts, mobility incidents (collisions, exploration with hands or feet, deviation from the path, and abrupt gait changes), and heart rate for the obstacle avoidance task, as well as success rate and mean normalized time for the object-finding task. The study design allows us to analyze which feedback modality users found more helpful, both subjectively and in terms of navigation performance.

Consistent with prior findings on ETAs, it is anticipated that both ETAs will decrease the number of mobility incidents and lower perceived workload, although walking speeds may decline due to learning effects. Given NOA’s camera-based system and spatialized auditory feedback, we expect it to provide broader obstacle coverage and more precise spatial awareness than the BuzzClip.

For the object-finding task, we anticipate both NOA configurations (AI- and DL-based versions) will guide participants effectively to target objects. We expect the DL version to result in faster task completion due to its concise spatial guidance, while the AI version will be preferred for its more descriptive and natural feedback.

## 2 Results

### 2.1 Obstacle avoidance task

Participants completed an obstacle avoidance task on a controlled indoor course under three conditions: using a white cane alone, using the cane with the BuzzClip, and using the cane with NOA. For each trial, we recorded mobility incidents, heart rate (HR), and the percentage of preferred walking speed (PPWS). After each condition, participants completed the NASA-TLX questionnaire and responded to a semi-structured interview at the end to provide feedback on their experience and perceived workload.

#### 2.1.1 Performance metrics

Repeated-measures ANOVA and Friedman tests revealed no difference in HR, deviation, changes in gait, or exploration with hands or feet across the three conditions. In contrast, a highly significant difference was found for the PPWS. Post-hoc comparisons showed that PPWS was significantly faster with the cane alone than with NOA (*t*(12) = 4.24, *p*uncorr = .001, pcorr = .002, *g* = 0.534) or the BuzzClip (*t*(12) = - 4.37, *p*uncorr = .001, *pcorr* = .002, *g* = - 0.695). A marginal increase between NOA and the BuzzClip was observed (*t*(12) = - 2.01, *p*uncorr = .067, *pcorr* = .067, *g* = - 0.173). A significant difference between the outward and return trials was noted, with faster PPWSs during the return trial (see Table S2). Cane-to-obstacle contact also differed significantly across conditions. Post-hoc tests highlighted significantly more contact when using the cane alone compared to with NOA (*W* = 3.00, *p*uncorr = .003, *pcorr* = .009, *g* = 0.996) and a non-significant increase when compared to the BuzzClip (*W* = 6.00, *p*uncorr = .051, *pcorr* = .076, *g* = - 0.649). Finally, results demonstrated a significant difference in body collisions with obstacles. Post-hoc comparisons highlighted significantly less body contact with NOA than with the cane alone (*W* = 6.00, *p*uncorr = .031, *pcorr* = .047, *g* = 0.608) or with the BuzzClip (*W* = 7.00, *p*uncorr = .019, *pcorr* = .047, *g* = 0.920) (see Fig. 2A). A complete overview of the omnibus tests can be found in Table 1.

**Figure 2.**
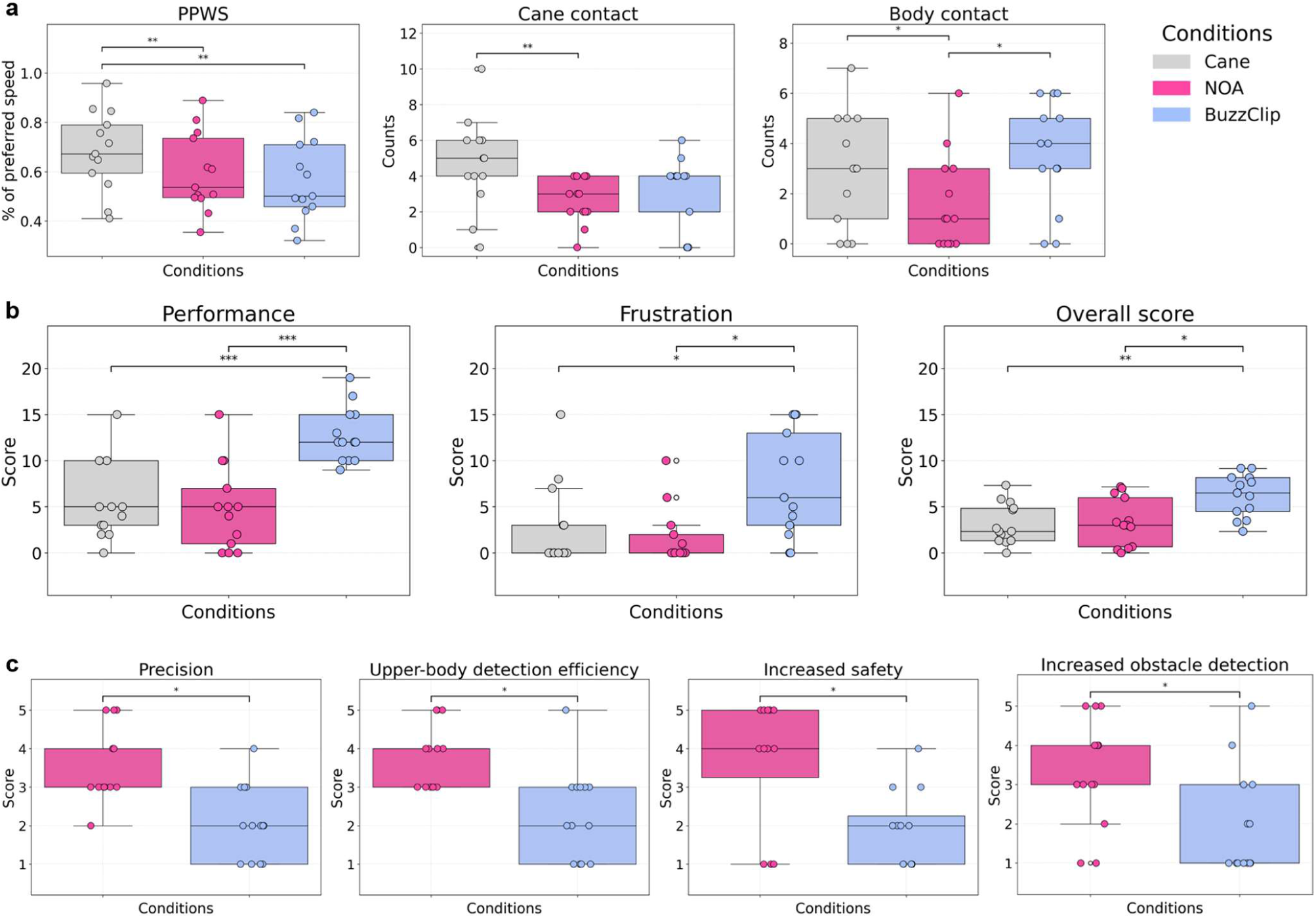
Scores across conditions during the obstacle avoidance task. (**a**) Performance metrics across conditions, including PPWS (%), cane contacts (counts), and body contacts (counts). (**b**) NASA-TLX workload scores (0–20), where lower scores indicate lower perceived workload. (**c**) Final questionnaire results, with scores ranging from 1 to 5. Bars with asterisks indicate statistically significant differences (*p* < 0.05: *, *p* < 0.01: **, *p* < 0.001: ***), based on FDR-corrected p-values.

**Table 1.** Performance metrics and NASA-TLX scores in the obstacle detection task. Repeated measures ANOVA tests (A) or Friedman tests (Fr) were performed depending on the normality of the data. F-values for ANOVA and *χ^2^*values for Friedman tests are reported. Values are presented as mean ± standard deviation (M ± SD). PPWS is expressed in percentage (%). All other performance metrics are event occurrences except heart rate, measured in beats per minute. NASA-TLX scores range from 0 to 20, with lower scores indicating lower workload. Effect sizes are generalized eta-squared *η^2^G* for ANOVA and Kendall’s *W* for Friedman tests. *P*-values are reported both uncorrected (*puncorr*) and corrected for multiple comparisons using the Benjamini–Hochberg procedure (*pcorr*).

| Condition | Cane | NOA | BuzzClip | Test type | Statistic | $p_{uncorr}$ | $p_{corr}$ | Effect size |
| --- | --- | --- | --- | --- | --- | --- | --- | --- |
| Performance metrics |  |  |  |  |  |  |  |  |
| PPWS (%) | 0.68 ± 0.16 | 0.60 ± 0.16 | 0.57 ± 0.17 | A | $F(2,24) = 16.45$ | **<0.001\*\*\*** | **0.002\*\*** | 0.093 |
| Cane contact | 4.69 ± 2.56 | 2.62 ± 1.26 | 3.15 ± 1.99 | Fr | $\chi^2(2) = 11.49$ | **0.003\*\*** | **0.011\*** | 0.442 |
| Body contact | 2.92 ± 2.25 | 1.62 ± 1.89 | 3.54 ± 2.15 | Fr | $\chi^2(2) = 9.26$ | **0.010\*** | **0.023\*** | 0.356 |
| Exploration | 0.46 ± 0.97 | 0.15 ± 0.38 | 0.46 ± 0.97 | Fr | $\chi^2(2) = 0.70$ | 0.705 | 0.705 | 0.027 |
| Gait change | 2.85 ± 2.03 | 2.85 ± 2.64 | 3.46 ± 2.93 | Fr | $\chi^2(2) = 5.45$ | 0.066 | 0.115 | 0.210 |
| Deviation | 0.08 ± 0.28 | 0.23 ± 0.60 | 0.00 ± 0.00 | Fr | $\chi^2(2) = 2.00$ | 0.368 | 0.429 | 0.077 |
| Heart rate (BPM) | 85.22 ± 11.85 | 80.43 ± 17.12 | 83.70 ± 10.93 | A | $F(2,24) = 1.30$ | 0.293 | 0.403 | 0.023 |
| NASA-TLX |  |  |  |  |  |  |  |  |
| Mental demand | 3.62 ± 3.43 | 7.46 ± 6.29 | 7.62 ± 5.42 | A | $F(2,24) = 2.78$ | 0.082 | 0.144 | 0.121 |
| Physical demand | 2.08 ± 2.14 | 1.62 ± 2.10 | 2.38 ± 2.36 | Fr | $\chi^2(2) = 2.10$ | 0.350 | 0.408 | 0.081 |
| Temporal demand | 2.46 ± 2.60 | 2.00 ± 3.27 | 2.54 ± 2.96 | Fr | $\chi^2(2) = 0.97$ | 0.616 | 0.616 | 0.037 |
| Performance | 5.69 ± 4.29 | 4.92 ± 4.61 | 12.77 ± 2.98 | A | $F(2,24) = 18.07$ | **<0.001\*\*\*** | **<0.001\*\*\*** | 0.455 |
| Effort | 2.46 ± 2.50 | 2.54 ± 3.13 | 4.46 ± 4.31 | Fr | $\chi^2(2) = 2.23$ | 0.327 | 0.408 | 0.086 |
| Frustration | 2.77 ± 4.62 | 1.69 ± 3.07 | 7.54 ± 5.74 | Fr | $\chi^2(2) = 11.76$ | **0.003\*\*** | **0.007\*\*** | 0.452 |
| Overall score | 3.18 ± 2.21 | 3.37 ± 2.60 | 6.22 ± 2.31 | A | $F(2,24) = 9.19$ | **0.001\*\*** | **0.004\*\*** | 0.270 |

#### 2.1.2 NASA-TLX

Similarly to the performance metrics, a repeated-measures ANOVA or Friedman test, followed by post-hoc comparisons were performed for each dimension of the NASA-TLX tool, as well as for the unweighted overall score (see Table 1 for a summary of results). No significant difference in reported mental, physical, temporal demand, or effort was found. However, a strong difference in perceived performance was demonstrated. The BuzzClip condition scored significantly higher (i.e., lower performance) than the cane alone (*t*(12) = 6.20, *p*uncorr = <.001, *pcorr* < .001, *g* = 1.856,) or with NOA (*t*(12) = 5.36, *p*uncorr = <.001, *pcorr* < .001, *g* = 1.959). Additionally, frustration scores differed significantly. The BuzzClip demonstrated higher scores than the cane alone (*W* = 7.00, *p*uncorr = .016, *pcorr* = .026, *g* = 0.886) or NOA (*W* = 6.00, *p*uncorr = .014, *p*corr = .026, *g* = 1.231). Results for the overall NASA-TLX score highlighted similar findings with significant differences in the repeated-measures ANOVA, and a higher score for the BuzzClip than the cane alone (t(12) = 3.67, *p*uncorr = .003, pcorr = .010, *g* = 1.301) or NOA (*t*(12) = 3.03, *p*uncorr = .011, *pcorr* = .016, *g* = 1.121) (see Fig. 2B).

#### 2.1.3 Final questionnaire

##### Likert-scale ratings

Depending on normality, paired t-tests or Wilcoxon signed rank tests were conducted for the Likert-scaled questions of the final questionnaire (see Table 2 for detailed results). Several significant differences were observed between the two devices. NOA was rated as significantly more precise in its obstacle localization capabilities than the BuzzClip. Similarly, participants found NOA better than the BuzzClip at detecting upper-body-level obstacles. For floor-level obstacles, the difference was significant when uncorrected but not after FDR correction; however, the rank biserial correlation was very strong. This suggests a robust practical difference, despite limited statistical power. When comparing each device to the cane-alone condition, participants expressed a significantly larger perceived increase in safety, and obstacle detection with NOA than with the BuzzClip (see Fig. 2C). Regarding trust during navigation, although the corrected p-value exceeded the conventional significance threshold, the relatively high Cohen’s d suggests a potential trend toward greater perceived trust with NOA compared to the BuzzClip. When questioned about the initial training phase, ease of learning tended to be higher for the BuzzClip. Although the difference did not reach statistical significance, a medium effect size was observed. No trend or significant difference was found in reported ease of use, comfort, or stress. Similarly, adequate learning time and quantity of information provided by the device did not differ significantly between devices.

**Table 2.** Responses to Likert-scale questions in the obstacle detection task. T-tests (T) or Wilcoxon signed-rank tests (W) were performed depending on the normality of the data. t-values for paired t-tests and *W* values for Wilcoxon signed-rank tests are reported. Values are presented as mean ± standard deviation (M ± SD). Scores range from 1 to 5. For adequate learning time and quantity of information: 1 = too little, 3 = ideal, 5 = too much. Effect sizes are Cohen’s *d* for t-tests and rank biserial correlation *rrb* for Wilcoxon tests. *P*-values are reported both uncorrected (*puncorr*) and corrected for multiple comparisons using the Benjamini–Hochberg procedure (*pcorr*).

| Condition | NOA | BuzzClip | Test type | Statistic | $p_{uncorr}$ | $p_{corr}$ | Effect size |
| --- | --- | --- | --- | --- | --- | --- | --- |
| Ease of learning | 3.77 ± 1.17 | 4.54 ± 0.88 | T | $t(12) = -1.95$ | 0.075 | 0.139 | 0.746 |
| Adequate learning time | 2.92 ± 0.64 | 3.31 ± 0.75 | W | $W = 0.00$ | 0.250 | 0.361 | -1.000 |
| Ease of use | 4.00 ± 0.91 | 4.08 ± 1.32 | T | $t(12) = -0.16$ | 0.874 | 0.874 | 0.068 |
| Comfort | 3.77 ± 0.93 | 4.38 ± 1.33 | W | $W = 18.50$ | 0.203 | 0.330 | -0.439 |
| Cane use interference | 1.31 ± 0.63 | 1.62 ± 1.04 | W | $W = 1.00$ | 0.500 | 0.591 | -0.667 |
| Increased trust | 3.54 ± 1.56 | 2.38 ± 1.26 | T | $t(12) = 2.56$ | **0.025\*** | 0.065 | 0.813 |
| Increased safety | 3.67 ± 1.67 | 1.92 ± 1.00 | W | $W = 0.00$ | **0.008\*\*** | **0.025\*** | 1.000 |
| Increased obstacle detection | 3.31 ± 1.38 | 2.00 ± 1.35 | W | $W = 0.00$ | **0.008\*\*** | **0.025\*** | 1.000 |
| Floor-level detection efficiency | 3.54 ± 1.27 | 2.23 ± 1.01 | W | $W = 1.50$ | **0.047\*** | 0.102 | 0.893 |
| Upper-body detection efficiency | 3.77 ± 0.83 | 2.31 ± 1.18 | T | $t(12) = 3.96$ | **0.002\*\*** | **0.022\*** | 1.430 |
| Stress level | 1.46 ± 0.78 | 1.62 ± 1.19 | W | $W = 3.50$ | 0.750 | 0.812 | -0.300 |
| Adequate quantity of information | 3.15 ± 0.90 | 2.77 ± 1.30 | T | $t(12) = 0.86$ | 0.406 | 0.527 | 0.344 |
| Precision | 3.54 ± 0.97 | 2.08 ± 0.95 | T | $t(12) = 3.63$ | **0.003\*\*** | **0.022\*** | 1.521 |

##### Device preferences

Participants were asked three questions per device and had to answer by yes, no, or undecided. To compare participants’ answers between the NOA and the BuzzClip, McNemar’s tests were conducted with responses collapsed into two categories (“yes” and “no” + “undecided“). A detailed summary of results can be found in Table S3. A significantly greater proportion of participants considered NOA to be a good complement to the white cane compared to the BuzzClip (100% vs. 53.9%). Similarly, more participants felt that NOA would increase their mobility on unfamiliar routes (76.9% vs. 23.1%). No significant difference was observed for mobility on familiar routes (61.5% vs. 38.5%).

A Chi-square goodness-of-fit test was conducted to examine participants’ preferences across the three options (NOA / BuzzClip / neither). Among the 13 participants, 12 preferred NOA, none preferred the BuzzClip, and one expressed no preference for either device. This distribution significantly deviated from a uniform distribution (χ^2^(2) = 20.46, *p* = < .001, *w* = 1.255).

##### Open-ended responses

Participants were asked whether they experienced any difficulties during the learning phase with both devices. Out of the 13 participants, 6 did not experience any difficulty with NOA, while 4 of them had trouble understanding the obstacle detection system, in particular the spatialization of the warning sounds. Moreover, 3 participants shared that despite not being required to set up the device themselves for the study, the user manual and settings would be difficult to understand if they had to do so autonomously. Concerning the BuzzClip, 7 participants did not highlight any difficult aspects. However, 4 of them noted that the vibrations were too weak, making them hard to discern from the natural vibrations coming from the white cane’s tip, and 2 participants found the system not precise enough.

Several recurring advantages of NOA were expressed. The majority of participants appreciated the precision of the spatialized sounds, particularly the device’s capacity to detect and distinguish obstacles at upper-body level (*n* = 8). 4 participants found the audio cues easy to interpret. Additionally, 4 participants valued the AI functionalities, and 3 appreciated NOA’s flexibility and all-in-one nature. Three shared that the device provided an added sense of safety and independence. However, some disadvantages were also reported, most notably the weight and size of the device (*n* = 8). They described the device as too heavy or bulky, particularly when wearing light clothing or backpacks. Despite the general appreciation for the obstacle detection system, 4 participants noted adoption challenges, including that the sound cues were too frequent, too loud, or had pitch variations that were difficult to interpret. Some also reported that obstacle alerts either arrived too late or failed to trigger altogether. Two participants did not report any disadvantages.

The BuzzClip was mainly praised for its small size and light weight design (n = 7), as well as its simplicity (*n* = 3). One participant mentioned its potential utility for deaf-blind individuals and children. However, 3 participants did not identify any advantages. Several drawbacks were highlighted, in particular the low intensity (*n* = 6) and high recurrence (*n* = 5) of vibrations, which some found confusing or distracting. Five participants considered the system insufficiently precise, citing a lack of information regarding obstacle elevation and unknown field of view. One participant did not report any disadvantages. A summary of all participants’ responses is provided in Table S4.

### 2.2 Object-finding task

In the second task, participants were asked to locate a person in a room based on auditory instructions provided by the NOA device with its object-finding functionality. Two different versions of the functionality were tested by participants: one using an AI-based software and the other using a DL-based version. The obstacle avoidance feature studied in the first task was not activated during this task. The task was performed under two conditions: one using an AI-based version and the other using a deep learning version of the object-finding functionality. Each participant completed three trials per condition. Normalized mean time and success rate were recorded. As in the first task, participants completed the NASA-TLX questionnaire and responded to a semi-structured interview to provide feedback on their experience and perceived workload.

#### 2.2.1 Performance Metrics

Paired t-tests were performed to compare normalized mean time and success rate. No significant difference in normalized mean time was observed (AI: *M* = 3.78, *SD* = 1.48; DL: *M* = 4.10, *SD* = 1.47; *t*(20) = - 0.54, *p*uncorr = .600, *pcorr* = .600, *d* = 0.216). Success rates demonstrated a trend, with the DL version outperforming the AI version, corresponding to a large effect size (*d* = 0.891), indicating a meaningful performance advantage, independent of statistical significance after correction (AI: *M* = 0.62, *SD* = 0.18; DL: *M* = 0.79, *SD* = 0.22; *t*(20) = - 2.21, *p*uncorr = .047, *pcorr* = .094).

#### 2.2.2 NASA-TLX

Depending on normality, paired t-tests or Wilcoxon signed-rank tests were performed on the six dimensions and overall score of the NASA-TLX. Results can be found in Table 3. No significant difference in reported mental and physical demand or frustration were observed. However, temporal demand differed significantly, with the DL version scoring lower (i.e., less demand) than the AI version. Similarly, significant differences were observed in effort and perceived performance. Finally, the DL version yielded a significantly lower overall reported workload score than the AI version (see Fig. 3A).

**Figure 3.**
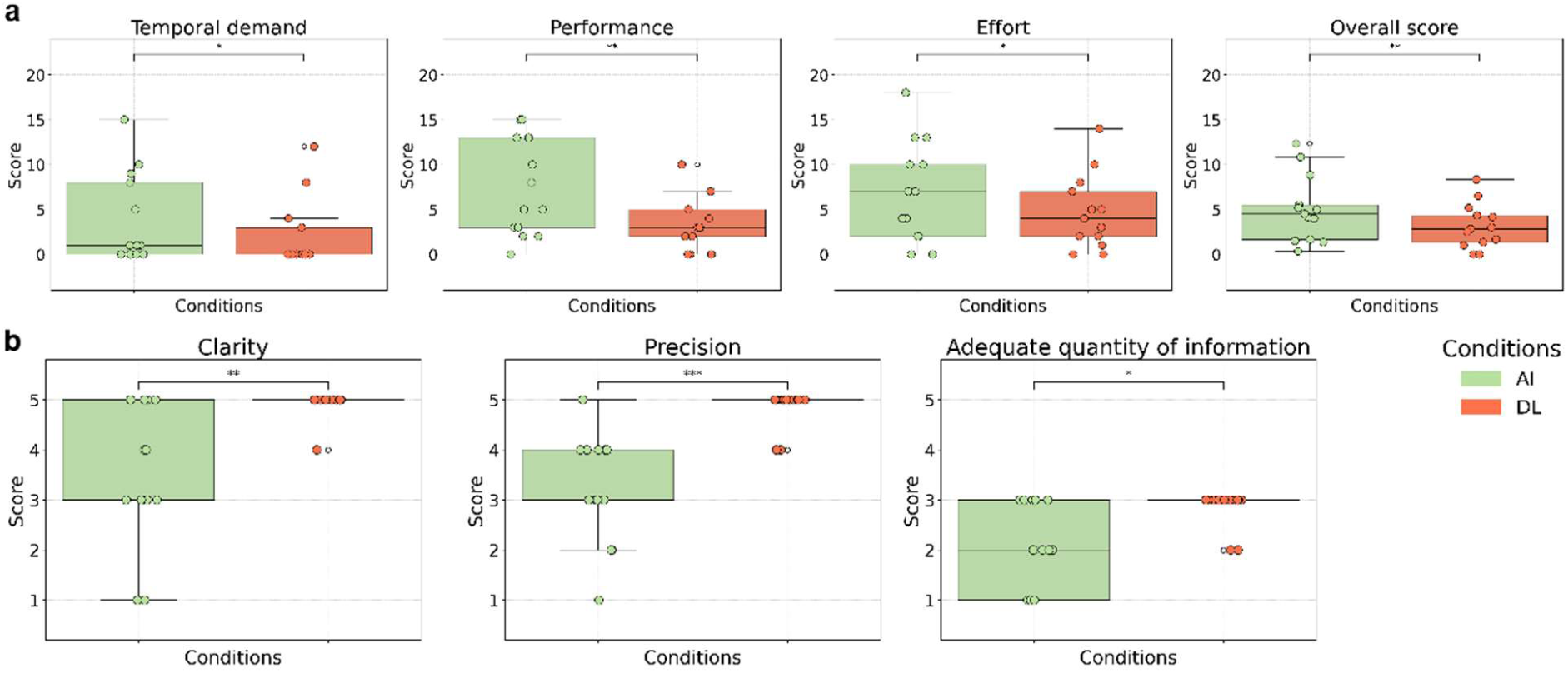
Scores across both conditions during the object-finding task. (**a**) NASA-TLX workload scores (0-20), where lower scores indicated lower perceived workload. (**b**) Final questionnaire results, with scores ranging from 1 to 5. For adequate quantity of information: 1 = too little, 3 = ideal, 5 = too much. Bars with asterisks indicate statistically significant differences (*p* < 0.05: *, *p* < 0.01: **, *p* < 0.001: ***), based on FDR-corrected p-values.

**Table 3.** NASA-TLX scores and final questionnaire results in the object-finding task. . T-tests (T) or Wilcoxon signed-rank tests (W) were performed depending on the normality of the data. t-values for paired t-tests and *W* values for Wilcoxon signed-rank tests are reported. Values are presented as mean ± standard deviation (M ± SD). NASA-TLX scores range from 0 to 20, with lower scores indicating lower workload. Final questionnaire scores range from 1 to 5. For adequate quantity of information: 1 = too little, 3 = ideal, 5 = too much. Effect sizes are Cohen’s *d* for t-tests and rank biserial correlation *rrb* for Wilcoxon tests. *P*-values are reported both uncorrected (*puncorr*) and corrected for multiple comparisons using the Benjamini–Hochberg procedure (*pcorr*).

| Condition | AI | DL | Test type | Statistic | *p<sub>uncorr</sub>* | *p<sub>corr</sub>* | Effect size |
| --- | --- | --- | --- | --- | --- | --- | --- |
| NASA-TLX |  |  |  |  |  |  |  |
| Mental demand | 6.54 ± 5.30 | 5.31 ± 5.68 | T | *t*(12) = 1.10 | 0.292 | 0.292 | 0.224 |
| Physical demand | 2.92 ± 3.90 | 2.08 ± 2.96 | W | *W* = 1.00 | 0.250 | 0.292 | 0.800 |
| Temporal demand | 3.85 ± 5.05 | 2.08 ± 3.84 | W | *W* = 0.00 | **0.008\*\*** | **0.018\*** | 1.000 |
| Performance | 7.23 ± 5.39 | 3.38 ± 2.90 | T | *t*(12) = 3.76 | **0.003\*\*** | **0.010\*\*** | 0.889 |
| Effort | 6.92 ± 5.60 | 4.69 ± 4.15 | W | *W* = 1.50 | **0.023\*** | **0.041\*** | 0.917 |
| Frustration | 2.62 ± 4.05 | 1.31 ± 2.10 | W | *W* = 0.00 | 0.125 | 0.175 | 1.000 |
| Overall score | 5.01 ± 3.69 | 3.14 ± 2.50 | T | *t*(12) = 3.98 | **0.002\*\*** | **0.010\*\*** | 0.594 |
| Final questionnaire |  |  |  |  |  |  |  |
| Complexity | 1.92 ± 1.32 | 1.15 ± 0.38 | W | *W* = 0.00 | 0.062 | 0.078 | 1.000 |
| Clarity | 3.46 ± 1.39 | 4.92 ± 0.28 | W | *W* = 0.00 | **0.004\*\*** | **0.010\*\*** | -1.000 |
| Stress level | 1.31 ± 0.75 | 1.15 ± 0.55 | W | *W* = 0.00 | 1.000 | 1.000 | 1.000 |
| Precision | 3.23 ± 1.09 | 4.85 ± 0.38 | T | *t*(12) = - 5.58 | **<0.001\*\*\*** | **<0.001\*\*\*** | 1.978 |
| Adequate quantity of information | 2.08 ± 0.86 | 2.85 ± 0.38 | W | *W* = 0.00 | **0.008\*** | **0.013\*** | -1.000 |

#### 2.2.3 Final questionnaire

Similarly to the NASA-TLX tool, questions from the final questionnaire were tested for normality before performing paired t-tests or Wilcoxon signed-rank tests accordingly. A significant difference in clarity was found between both versions, highlighting a higher score for the DL version. Similarly, the DL version’s instructions were considered more precise than the AI version and their quantity more adequate than the AI version, which scored lower (i.e., insufficient instructions) (see Fig. 3B). No significant difference was observed for stress level. A marginal trend was observed for task complexity, with the AI version perceived as slightly more complex. A complete summary of results can be found in Table 3).

A chi-square goodness-of-fit test was conducted to examine participants’ preferences between the DL version and AI version. All participants preferred the former, yielding a significant result (*χ^2^*(2) = 26.00, *p* < .001, *w* = 1.41).

## 3 Discussion

The present study examined the performance of BVI participants using two ETAs across an obstacle avoidance task and object-finding task.

The obstacle avoidance task was performed three times - with the cane alone, with the cane and NOA, and with the cane and the BuzzClip. It is important to clarify that the cane-alone condition was not intended as a direct competitor to the ETAs, but rather as a baseline to evaluate the added value and performance of NOA and the BuzzClip when used in conjunction with the cane. Quantitative results demonstrated that participants employed their cane less with NOA (i.e., less contacts with obstacles) and collided less with obstacles than with the BuzzClip or with the cane alone. These results indicate enhanced obstacle avoidance. Questionnaire responses echoed this: NOA received significantly higher ratings than the BuzzClip in precision, detection capabilities, safety, and upper-body-level obstacle detection. Several participants appreciated the accuracy of the spatialized sounds, which allowed them to locate obstacles efficiently. Moreover, despite its technological complexity noted by some participants, NOA did not lead to higher workload according to the NASA-TLX results. This finding is promising, as it demonstrates that ETAs can improve mobility and safety without creating cognitive overload. Indeed, increased workload and cognitive overload have been a concern in the development and research of ETAs [17–20]. The additional information to process can be demanding for users, which hinders the potential advantages of the devices. However, this was not the case with NOA, which could suggest that the cues are adequate and intuitive for adult BVI users.

On the other hand, the BuzzClip was praised for its light weight and simplicity, contrasting with the bulkiness of NOA noted by several participants. However, they reported poorer obstacle detection. Moreover, the BuzzClip scored more poorly on workload, specifically due to increased frustration and lower perceived performance. This was corroborated by the performance metrics that showed no improvements in mobility incidents when using the BuzzClip. One explanation for such poor outcome comes from the cues provided by the device. Notably, participants complained that the vibrations were often uninformative and confused with the cane feedback. This could potentially explain why the device felt more demanding and less efficient, even if it was not technically more complex than NOA.

Walking speed was slower with both assistive devices when compared to the cane alone. This is consistent with prior findings that suggest that reduced speeds could reflect the need to process unfamiliar cues [16,21]. This finding is also aligned with observations from the VibroSight tactile device study by Jin et al. [22], which used motion-tracking analysis to quantitatively assess mobility performance. In that study, participants, despite achieving effective obstacle detection and intuitive directional feedback after minimal training (about 20 minutes), exhibited slower walking speeds as they processed additional sensory information. This suggests that extended familiarization or training could help users integrate novel feedback more efficiently, potentially reducing such differences in speed<u>—an aspect that should be investigated</u> in future research. Furthermore, several studies have suggested a link between reduced walking speeds and generally increased cognitive workload during mobility tasks [23,24]. Interestingly, our results diverge partially from this theory: although walking speeds were similar across devices, only the BuzzClip was associated with a higher perceived cognitive workload, while that reported with NOA was not significantly different from that with the white cane alone. This suggests that participants may have slowed down for different reasons. Reduced speed for the BuzzClip might be at least partially a compensatory reaction to the higher cognitive demands, whereas the slowing when using NOA might instead be caused mainly by unfamiliarity-induced caution. Moreover, a key difference with the long cane is that NOA extends the localization space to greater distances, providing earlier access to spatial information. This broader detection range may require additional time to interpret, potentially contributing to the initially more cautious walking behavior.

Although non-significant results, the large effect size and participants’ feedback indicate a more demanding initial learning curve for NOA, particularly due to the complex 3D-sound system. However, this did not translate to differences in performance results or workload. This could imply that even though NOA might require more time to adapt, it becomes more intuitive with practice and usage. Nonetheless, simple devices like the BuzzClip may offer faster initial training, but their lower effectiveness may limit long-term usability.

When asked about potentially adopting one of the devices, all participants selected NOA, except for one who chose neither device. Everyone agreed that NOA could serve as a useful complement to the white cane. Only about half felt the same about the BuzzClip. Participants were also asked whether the devices might help them travel more frequently on familiar and unfamiliar routes. While no significant difference was observed for familiar routes, over 75% believed NOA could increase their mobility on unfamiliar routes, compared to only 23% for the BuzzClip. These results demonstrate high appreciation and potential adoption of NOA. It also suggests that NOA may be particularly helpful on unknown routes where the cane might not be sufficient when encountering new hazards, and where longer-distance detection could offer a meaningful advantage.

Overall, the present results highlight strengths and weaknesses in both devices. NOA was generally more appreciated and provided increased mobility. The BuzzClip was praised for its simplicity and light weight. While its design was valued, the feedback system lacked reliability and precision for potential adoption, yielding no mobility advantages and contributing to increased workload. Nonetheless, the BuzzClip highlights the desire for a simple, compact solution, contrasting with NOA’s bulkiness and complexity. These observations suggest that both devices could benefit from targeted improvements: ergonomic refinements for NOA, and enhanced feedback clarity for the BuzzClip. Future hardware and software developments may help address these aspects, supporting broader usability and adoption across different user preferences and contexts.

The results also raise the question of the best type of feedback to convey obstacle type and location efficiently and intuitively. This issue has been widely explored; previous research investigated auditory, tactile, and multisensory feedback systems. Yet, the findings are still conflicting, and no clear consensus has emerged on an ideal solution [16,18,25]. In our study, participants preferred NOA’s spatialized audio because it conveyed precise obstacle location, including elevation and direction cues. BuzzClip’s haptic feedback lacked this precision and did not provide any information on obstacle height. Participants also criticized the BuzzClip’s vibrations, describing them as too weak and too frequent, which could have caused further confusion. Nonetheless, the results suggest that spatialized audio feedback may offer advantages in terms of precision and intuitiveness. However, it is important to recognize that these devices differed not just in feedback modality, but in complexity and output richness. Therefore, preferences for one form of feedback over another cannot be entirely disentangled from the broader findings. Moreover, it is worth noting that our study was conducted in an indoor controlled environment lacking background noise from real-world settings. Despite NOA’s bone-conducting headphones, auditory feedback may still interfere with users’ perception of surrounding sounds, critical for the navigation and mobility of BVI individuals in complex environments or simply be masked by background noise. In this regard, haptic feedback may offer a natural advantage. No difference was observed in mean time between the AI and DL versions in the object-finding task. For success rate, the DL version showed a large effect size (*d* = 0.891), indicating a stronger advantage in performance, independent of statistical significance.

Participants’ feedback corroborates this proposition with a lower perceived workload with the DL version. Moreover, participants found the DL version clearer, more precise, and thought that the quantity of indications was more adequate, with the AI version not providing enough information to complete the task efficiently. Consequently, all the participants preferred the DL version. Several of them explained that they particularly appreciated the details provided about orientation and distance (e.g., “*Person to your left at 10 o’clock, 3 meters*“) while others found the spatialized sounds especially useful to locate the person. One participant noted that while they preferred the DL version for object-finding due to its precision, they felt that the richer, more descriptive language of the AI version might be more useful in other situations, such as general scene descriptions. These results suggest that precise initial indications, as well as continued instructions in the form of a spatialized sound, are appreciated and efficient for precise localization of an object or person. However, generative AI has the advantage of providing more details about a scene in a richer language which can be favored in other contexts, such as understanding one’s environment in a broader way. It may therefore be advantageous to have both options integrated into a device and used at an individual’s discretion depending on the context and their preferences.

While our study yields promising findings, several limitations must be acknowledged. First, our sample size (*n* = 13) may have restricted the statistical power and generalizability of the results. Although small sample groups are common among experimental studies on ETAs [18,21,23], future research should aim for larger cohorts. This would enable more robust conclusions. Moreover, it would allow for the implementation of user-profiling based on participants’ individual characteristics. For example, certain types of ETAs may be better suited to specific user profiles. However, identifying these patterns requires larger datasets, which was not possible in our study.

Second, we believe that, while indoor settings provide a controlled experimental environment, to truly study the impact of ETAs, trials should also be conducted outdoors. Despite being appropriate for indoor mobility, NOA and the BuzzClip are largely designed for outdoor travel and would therefore benefit from being tested outside. The presence of moving obstacles, environmental noise, or uneven ground textures may influence both device performance and user experience in ways not captured in the present study. In real-world contexts, the extended detection range of such devices could potentially lead to an overload of non-critical information that may distract or even endanger users. This limitation also applies to the object-finding task, as real-world environments introduce additional challenges such as dynamic targets or crowded scenes. A more realistic setting would not only challenge the software versions more effectively, but also help to determine whether one version is better suited to particular environments or use cases.

Third, our study deliberately focused on early-stage use. Participants spent about an hour at home studying both devices and about 45 minutes of guided training on the test day. While this duration was sufficient for basic comprehension of device cues, it does not reflect long-term or expert use. The unfamiliarity likely contributed to the slower walking speeds during testing. Subsequent studies should consider evaluating performance with experienced users or over extended adaptation periods to better capture sustained usability.

Overall, participants improved their mobility, while maintaining a reasonable cognitive workload and feeling safer with NOA than with the BuzzClip or white cane alone. Notably, 12 out of 13 participants indicated they would consider adopting the device. The results suggest strong potential for real-world implementation.

However, as stated above, further research is needed to validate these findings. In particular, future studies should investigate NOA’s performance during outdoor navigation and over longer training periods. Longitudinal studies would be particularly useful to assess continued engagement, trust, and real-life retention. Moreover, several design considerations can be drawn from our findings to guide future ETA development. First, a lightweight and ergonomic design is essential to ensure comfort and long-term wearability, as excessive bulk or weight can discourage adoption despite functional advantages. Second, feedback precision and intuitiveness are critical: our results highlight the benefits of NOA’s precise spatialized auditory cues for accurate obstacle avoidance and the limited spatial accuracy of the BuzzClip’s haptic feedback. This does not imply that one modality is inherently superior but rather that precision and accuracy are crucial to support users and prevent cognitive overload. Third, training and familiarization protocols should be systematically integrated into device onboarding, as our results show that new systems can be demanding and may slow down users. Finally, participants demonstrated enthusiasm for all-in-one devices like NOA, which provide personalization and context-dependent modes according to user goals. The object-finding task further highlighted interest in such functionalities, showing that both descriptive and concise feedback could be helpful depending on the users’ objectives. \Together, these insights support the creation of ETAs that are both user-centered and technically effective.

Finally, we note that NOA has been commercially available since 2024 and is already in use by early adopters worldwide. Our findings offer valuable insights for refining the device and evaluating its impact as it continues to scale.

## 4 Methods

Our study aimed to evaluate the performance and perceived usability of two ETAs, NOA and the BuzzClip, in enhancing mobility with the white cane. We employed a within-subject, counterbalanced design consisting of two tasks: an obstacle detection task and an object-localization task (Figs. 4 and 5). The obstacle detection task included three conditions: white cane alone, white cane and the BuzzClip, and white cane and NOA. The object-finding task compared two versions of NOA’s object-finding functionality.

**Figure 4.**
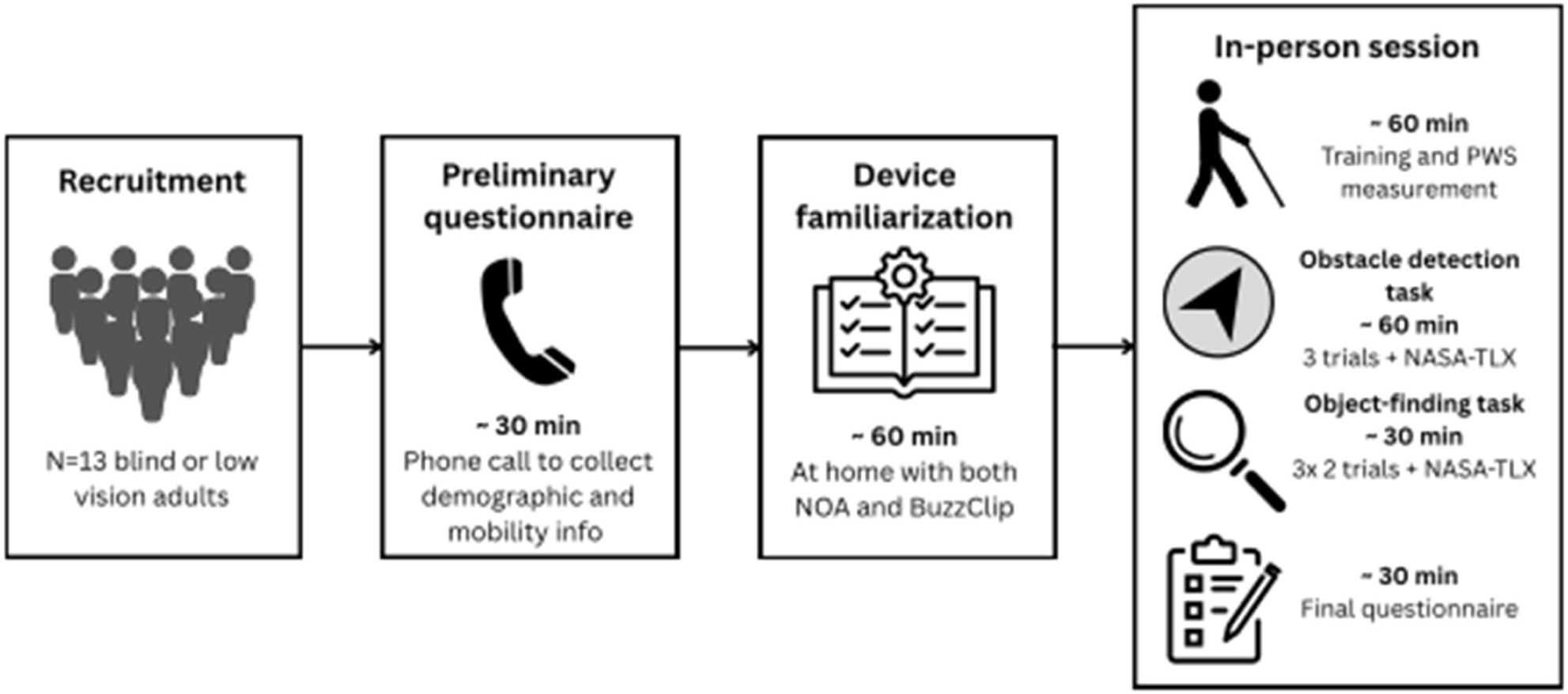
Overview of the study design and time course. Each participant (n=13) completed a preliminary phone questionnaire, at-home familiarization with the device manuals, and a single 3-hour in-person session involving obstacle detection and object-finding tasks.

**Figure 5.**
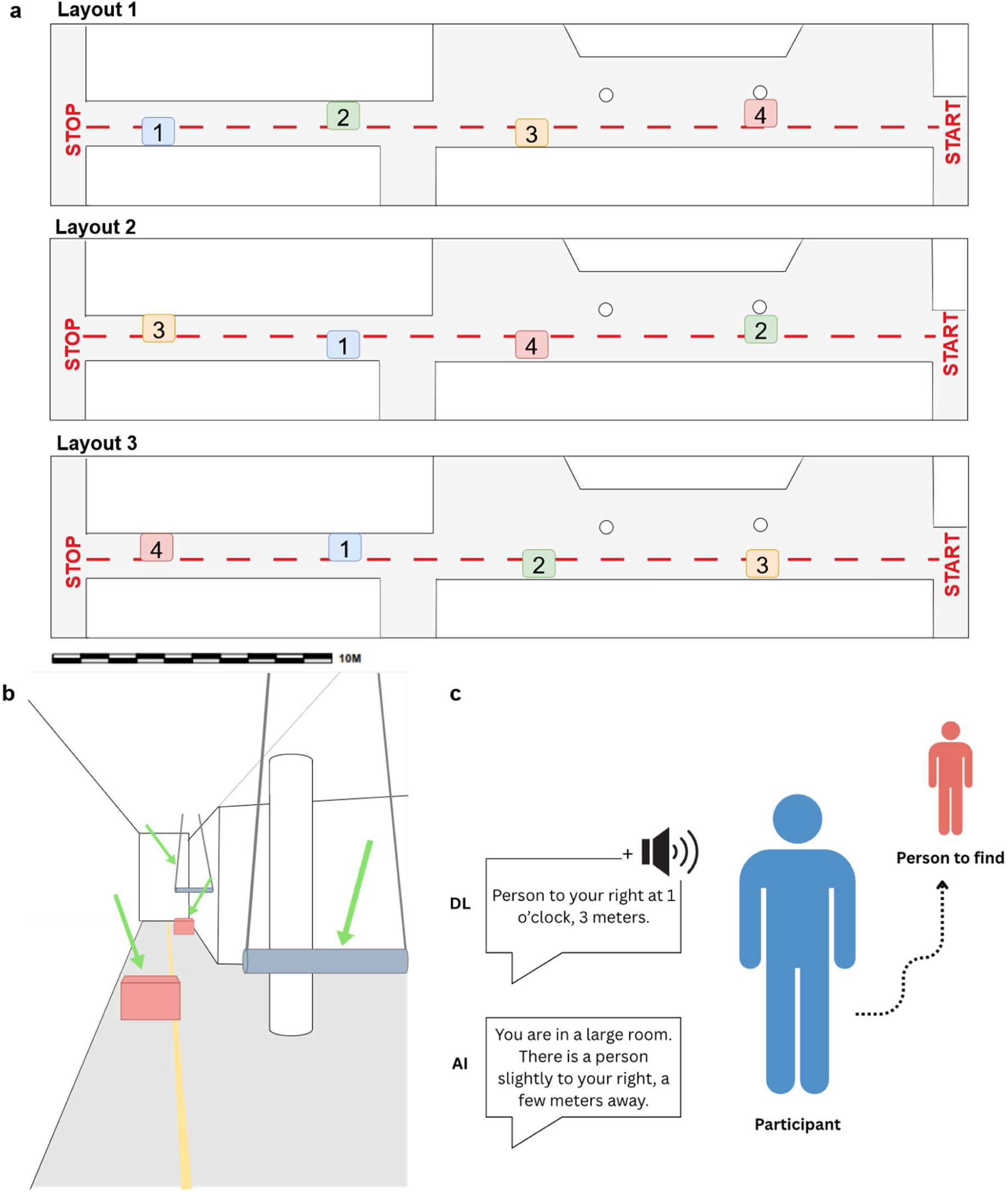
Experimental setups used in the study. (**a**) The three obstacle layouts used in the obstacle avoidance task. Obstacles 1 and 3 are cardboard boxes on the floor; obstacles 2 and 4 are foam rolls at waist and head level. (**b**) The 28-meter corridor setup for the obstacle avoidance task, with obstacles highlighted by green arrows and the guidance line in yellow. (**c**) The procedure of the object-finding task. The DL version provided a description of the target’s location and spatialized sounds. The AI version provided a description only.

To assess the ETAs’ performance and user experience, we collected a combination of quantitative performance metrics and qualitative data. Qualitative data included semi-structured interviews and ratings of perceived workload. All statistical analyses and comparisons were prespecified and are described in detail, below.

### 4.1 Participants

The study was approved by The Cantonal Ethical Commission for Research on Human Subjects (CER-VD) (protocol number 2024-02523), and all procedures complied with the current version of the Declaration of Helsinki and the ICH-GCP as well as all national legal and regulatory requirements.

The study involved 13 participants, including 8 males and 5 females, aged between 25 and 85 years (Mean = 55.77, SD = 19.78 years). Participants were BVI individuals with varying degrees of vision loss. Five reported having low vision, while eight were legally blind, with four of them reporting total blindness. Vision levels were self-identified by the participants and correspond to the definitions used in Switzerland (0.05 ≥ VA < 0.3 for low vision; VA < 0.05 for legal blindness; and no perception of light or shapes for total blindness). Participants were recruited through local organizations that support BVI. They were provided with a detailed explanation of the study’s purpose before obtaining informed consent. The following criteria were established for inclusion: The participants had to be 18 years old or older, be blind or visually impaired, and able travel outside independently. Participants were excluded if: they were regular BuzzClip or NOA user (at least once a week), had cognitive, additional motor or physical mobility impairments, had an incapacity of discernment or to understand instructions. Demographic information on the participants can be found in Table S5. Travel costs were reimbursed but participants did not receive any other financial compensation for their participation.

### 4.2 Materials

The study investigated two ETAs: the BuzzClip (iMerciv Inc., Toronto, Canada) and NOA (Biped Robotics SA, Epalinges, Switzerland) devices. The performance of both devices was evaluated through an obstacle avoidance task in a controlled indoor environment (see Fig. 5A-B). Additionally, a new object detection and guidance feature on the NOA device was also investigated (see Fig 5C).

#### 4.2.1 BuzzClip

The BuzzClip is a small wearable device weighing about 50 grams that can be clipped to clothing, attached to a cane, or used handheld (see Fig. 1A). It utilizes ultrasonic sensors to detect obstacles in the user’s path and offers feedback through vibrations to warn the user [26,27]. This approach is similar to other travel aids such as the “EyeCane” [28]. The intensity and frequency of the vibrations increase as the user approaches an obstacle, helping them situate the hazard. The sensors have a maximum angle coverage of 30° and a detection range from 1 to 3 meters. The device was designed as a complement to the white cane or to the guide dog, focusing on detecting upper body and head level obstacles. The details overviewed here are derived from the BuzzClip’s user manual [29].

Since the start of our study, the BuzzClip has been discontinued by its manufacturer. Nevertheless, due to its functional similarities with other current assistive technologies, we still considered it valuable and relevant to include the BuzzClip in our comparative evaluation.

#### 4.2.2 NOA

The NOA device is a mobility vest worn on the shoulders. It has three Intel RealSense D430 cameras on the left of the chest, a processing unit on the right, a battery at the back of the neck, and weighs about 1.3 kilograms (2.8 lbs) (see Fig. 1B). The cameras capture synchronized depth and 2D RGB images at 10 frames per second with a field of view of 90° vertically and 170° horizontally, and a range of 0.3-10 meters. The images are processed locally by the onboard CPU. NOA’s main functionality is obstacle avoidance, which was tested in the study’s first task. Ground detection is performed through 3D point classification, and obstacles are aggregated using agglomerative algorithms. Potential collisions are filtered using rule-based selection. When an obstacle is identified, audio feedback is generated: spatial orientation is conveyed via an head-related transfer function (HRTF-based) spatialization algorithm, vertical position via a pitch gradient, and distance via audio cue frequency. The HRTF spatialization algorithm was developed by GCRadix (https://www.gcradix.de). Feedback is transmitted via Bluetooth Low Energy (BLE) to Shokz OpenRun Pro bone-conducting headphones (Shokz, https://shokz.com).

NOA also integrates GPS navigation with turn-by-turn directions, as well as an object-finding feature, that leverages generative AI features to recognize and describe various landmarks (e.g., doors and exits, people, street crossings, bus stops). In a second task, the AI-based object-finding feature was tested and compared to a version based on a DL-based software. Details about this task are provided in Section 4.3.5.

Similarly to the BuzzClip, NOA is designed to be compatible with existing mobility aids, such as white canes or guide dogs [30,31].

### 4.3 Procedure

As shown in Fig. 4, the study included a preliminary questionnaire completed during a phone call, familiarization with the devices’ user manuals at home, and one individual in-person session per participant which lasted about 3 hours.

#### 4.3.1 Preliminary questionnaire

After receiving the consent from participants, a call was scheduled in the weeks ahead of the experiments. The purpose of the call was to complete a preliminary questionnaire covering demographic questions, condition, and mobility habits. Table S5 summarizes the key findings. Participants were informed that they would receive the user manuals for both devices one week prior to their scheduled appointment. All provided materials were official material from manufacturers. Both documents were sent in an accessible electronic and audio version. This step allowed the participants to familiarize themselves briefly with the devices beforehand. Participants were not required to spend a specific amount of time reviewing the materials but were asked to have a basic understanding of both devices prior to their session. On average, participants reported spending about one hour on this task.

#### 4.3.2 Training

On the day of the practical tests, the session started with a familiarization phase where participants were introduced to the devices in a calm room. This initial presentation was relatively brief, as participants had reported reviewing the manuals in advance. Following this, participants had the opportunity to practice using the devices in an open space where a few low-risk obstacles were placed around them. The training was considered complete when participants demonstrated a clear understanding of how to use the devices and could detect and locate an obstacle at a distance of at least 1.5 meters with both devices.

#### 4.3.3 Preferred walking speed

Before starting with the practical task, the preferred walking speed (PWS) of the participants was measured. The PWS is a measure of an individual’s optimal walking speed. It allows the measurement of walking efficiency, as the actual speed that the participants adopt during the practical tasks can be expressed as a percentage of their preferred walking speed (PPWS) [32–34]. The participants walked an unfamiliar 10-meter-long indoor hall in a calm, controlled environment. To measure the PWS, the sighted guide technique was used, where the participant is accompanied by a sighted guide [35]. Four trials were performed and the mean pace from the last three of these trials was used as the PWS; the first one being excluded to avoid any bias due to adaptation to the route itself. The average PWS across participants was 0.98.m/s (SD = 0.18.m/s).

#### 4.3.4 Obstacle avoidance task

After the training and PWS measurement, participants completed the obstacle avoidance task. They navigated an unfamiliar indoor route in a calm, controlled environment. The route consisted of a 28-meter straight indoor corridor with four obstacles made from materials designed to minimize injury in case of a collision (see Fig. 5B). Two obstacles at upper-body-level were foam rolls suspended from the ceiling by strings attached with magnetic hooks. These obstacles could move or detach upon impact to reduce injury. The two floor-level obstacles were empty cardboard boxes. Two research investigators were present during the task, with one responsible for intervening if a participant was at risk of injury. A straight line on the floor served as a sensory guide for participants. This line consisted of a 28-meter, 8 mm-diameter cord securely taped to the middle of the corridor. Similar to tactile paving found in public spaces, it provided participants with a navigational reference.

Participants were required to navigate the route while avoiding as many obstacles as possible. They completed the task under three conditions: (1) using only a white cane, (2) using a white cane with NOA, and (3) using a white cane with the BuzzClip. For each condition, participants traversed the corridor in both directions, with a different obstacle arrangement for each condition (see Fig. 5A). To control for learning and order effects, participants were assigned to one of three counterbalanced condition orders: (1) Cane → NOA → BuzzClip, (2) NOA → BuzzClip → Cane, and (3) BuzzClip → Cane → NOA.

While one investigator rearranged the obstacles, the second completed the NASA-TLX questionnaire with the participant. The NASA-TLX is a widely recognized tool consisting of six questions designed to evaluate the perceived workload associated with a task [36]. It was completed after each condition to avoid any cumulative effect between the three trials. Moreover, this ensured that the participant could not hear the sounds of obstacles being moved, which might have otherwise provided clues about the course layout.

Several metrics were measured during the obstacle avoidance task by one of the investigators: PPWS, cane to obstacle contact, HR, and mobility “incidents” (i.e., body-obstacle contact, exploration of surroundings with hands and feet, loss of balance, marked changes in walking speed, deviation from path).

The indoor mobility course and the performance metrics collected during the tasks were partially designed based on the framework of [37], who conducted an in-depth review of key factors in assessing indoor mobility performance.

#### 4.3.5 Object-finding task

Participants then proceeded with the “object-finding” task. This task involved searching for a person in a room by following auditory instructions provided by NOA. Two versions were used in this task:

1. **AI version**: The first version uses generative AI. The images captured by the device are transmitted from the local CPU to a cloud-based inference engine running Gemini Flash 2.0. Mobility-oriented prompts are dynamically selected based on the task demanded. The resulting text-to-speech (TTS) output is streamed via BLE to the user’s headphones. All data are permanently deleted from the cloud immediately after processing. This version provides a general but more natural description of the person’s location (e.g., “*You are in a room. There is a person slightly to your left, a few steps away*”). No additional information is provided following this instruction.
2. **DL version**: The second version applies a DL algorithm locally. It uses a trained semantic segmentation model to detect and recognize specific objects, running on the local Neural Processing Unit (NPU) at 4 frames per second. Feedback is streamed via BLE. The DL version offers a more specific description with both orientation and distance information (e.g., “*Person to your left at 11 o’clock, 3 meters*”). Following this first instruction, a 3D-spatialized sound (i.e., a “beep” sound) indicates the location of the target person throughout the search. During the task, the sound was present as long as the target person was present in the cameras’ field of view (170°) and was interrupted by the instructor at the end of the trial.

The task was conducted in an empty room measuring 5 meters by 5.5 meters, which was dimly lit (see Fig. 5C). Participants were instructed to keep their eyes closed if they had low vision (*n* = 7). No participants reported perceiving visual information during the task. During each trial, participants were placed on one side of the room, facing the rest of the space. The person they were instructed to find was positioned at a distance of 2 or 3 meters away from them and located at one of five possible clock-face positions: 10, 11, 12, 1, or 2 o’clock. Participants were told to listen to the initial auditory description of the person’s location and then begin walking toward that position. Once they thought that they were close enough to the person, they were permitted to use their hands to confirm whether the person was indeed in front of them.

Each participant completed the task three times per condition, with the order of conditions counterbalanced across participants. The task time and success rate were recorded for each trial. A trial was considered failed if the participant made more than three changes in direction, was unable to correctly identify the location of the person, abandoned the task, or took longer than 30 seconds to find the person. Additionally, the NASA-TLX questionnaire was performed for both versions.

#### 4.3.6 Final questionnaire

After completing the task, participants answered the last semi-structured questionnaire. It consisted of questions about the device training and usage, both tasks, and open questions.

### 4.4 Data analysis

Statistical analyses were performed in Python (v3.10.12) using the Statsmodels, SciPy, and Pingouin packages. Multiple comparisons were corrected using the Benjamini-Hochberg False Discovery Rate (FDR-BH) procedure. All analyses employed a significance threshold of (α = 0.05).

#### 4.4.1 Obstacle avoidance task

A repeated-measures analysis was conducted for the performance metrics as well as for the NASA-TLX tool. These included:

1. **Performance metrics**: PPWS, cane-to-obstacle contacts, body-to-obstacle contacts, exploration with hands and feet, deviations from the path, marked changes in walking speed, loss of balance, and HR. Loss of balance was removed from the analyses as no participant demonstrated any sign of such incident.
2. **NASA-TLX (0–20 scale)**: mental demand, physical demand, temporal demand, effort, performance, frustration, and overall score (mean of the 6 dimensions).

Possible differences in performance between the outward and return trial were investigated. To this matter, normality was verified using a Shapiro-Wilk test and QQ plots. Based on the outcome, a paired t-test, if normality was met, or a Wilcoxon signed-rank test, was conducted between both “rounds” for each performance metric.

After analyzing differences between both trials, the sum of both trials was computed for the cane-to-obstacle contacts and mobility incidents. For HR and PPWS, a mean of both trials was preferred. For each metric, normality was investigated, and, if met for all three conditions, a repeated-measures ANOVA was conducted, corrected with Greenhouse-Geisser if sphericity was violated. If normality was not met, a Friedman test was conducted instead. Finally, significant metrics were investigated with post-hoc pairwise comparisons.

The final questionnaire was comprised of four types of questions, each answered separately for NOA and the BuzzClip:

1. **Likert-scale questions (1-5)**: ease of learning, adequate learning time (1 = too little, 3 = ideal, 5 = too much), ease of use, comfort, interference in cane use, increased trust/safety/obstacle detection in comparison to the white cane alone, efficiency in detecting obstacles at upper-body/floor level, stress, adequate quantity of information provided by device (1 = too little, 3 = ideal, 5 = too much), and precision.
2. **Ternary-choice questions (Yes / No / Undecided)**: whether the devices were considered a good complement to the cane, whether they could increase their ability to travel on familiar and unfamiliar paths.
3. **Preference (NOA / BuzzClip / none)**: which device they would consider adopting in daily life.
4. **Open questions**: difficulties during the learning phase, advantages, and disadvantages of both devices.

For the Likert-scaled questions, if normality was achieved, paired t-tests were conducted to analyze potential differences between NOA and the BuzzClip. Else, Wilcoxon signed-rank tests were performed.

Participants’ preferences were analyzed using a Chi-square goodness-of-fit test. The ternary-choice questions were analyzed using McNemar’s tests. For this analysis, responses were collapsed into a binary outcome (’Yes’ versus ’No’ or ’Undecided’) to align with our primary interest in evaluating the practical relevance of each device, and potential differences between both. While this simplification sacrificed some ordinal nuance, it enhanced the robustness of statistical inference given the limited sample size and provided a clearer indication of the relative impact each device may have on participants’ mobility and independence.

Open questions were reviewed qualitatively to identify general patterns and recurring themes.

#### 4.4.2 Object-finding task

The following metrics were analyzed:

1. **Performance metrics**: normalized mean time, success rate. Trial times were normalized based on the distance to the target (i.e., 2 or 3 meters). The normalized mean times were computed, excluding failed trial times, as well as the success rate.
2. **NASA-TLX (0-20 scale)**: mental demand, physical demand, temporal demand, effort, performance, frustration, and overall score (mean of the 6 measures).
3. **Final questionnaire (Likert-scale, 1-5 unless specified)**: precision, clarity, difficulty, adequate quantity of information (1 = too little, 3 = ideal, 5 = too much), stress, and preferred version (AI / DL / None).

For each metric, normality was tested using a Shapiro-Wilk test and QQ plots. Based on the outcome, we applied a paired t-test if normality was met. Otherwise, a Wilcoxon signed-rank test was used. These comparisons were done for each metric between the two experimental conditions. To analyze participants’ preferences, a Chi-square goodness-of-fit test was performed.

## 5 Supplementary Materials

Tables S1 to S5

## 6 Data availability

Data and code used in the analysis are available from the corresponding authors upon request.

## Data Availability

All data produced in the present study are available upon reasonable request to the authors

## Acknowledgments

We thank Dr. Uta R. Roentgen and Ms. Fatima Anaflous for sharing their generous advice and expertise in planning the mobility course and refining the study tasks and questions. We thank all the participants for their engagement in the study. We also thank our colleague, Dr. Matthew Vowels, for his valuable support throughout the research process, particularly for his guidance during the data analysis phase.

## 7 Author contributions

Conceptualization: CEP, MMM

Data curation: CEP

Formal analysis: CEP, EVO

Funding acquisition: MMM, MF

Investigation: CEP, MF

Methodology: CEP, MMM

Project administration: CEP, MMM

Resources: CEP, MF, MMM

Software: MF

Supervision: MMM

Validation: MMM, EVO, MTW, MG

Visualization: CEP, EVO, MMM

Writing – original draft: CEP, MMM, EVO

Writing – review & editing: CEP, MMM, EVO, MTW, MG

## 8 Additional information

C. E. Pittet performed a research internship at Biped Robotics SA in the context of her MSc studies. M. Fabien is founder and CEO of Biped Robotics SA. While Biped Robotics provided funding, instrumentation, and software for this study, they were not involved in the study design, data analysis, or interpretation of results. No other co-author has any conflicts of interest with regard to this work.

---

**Table S1.**
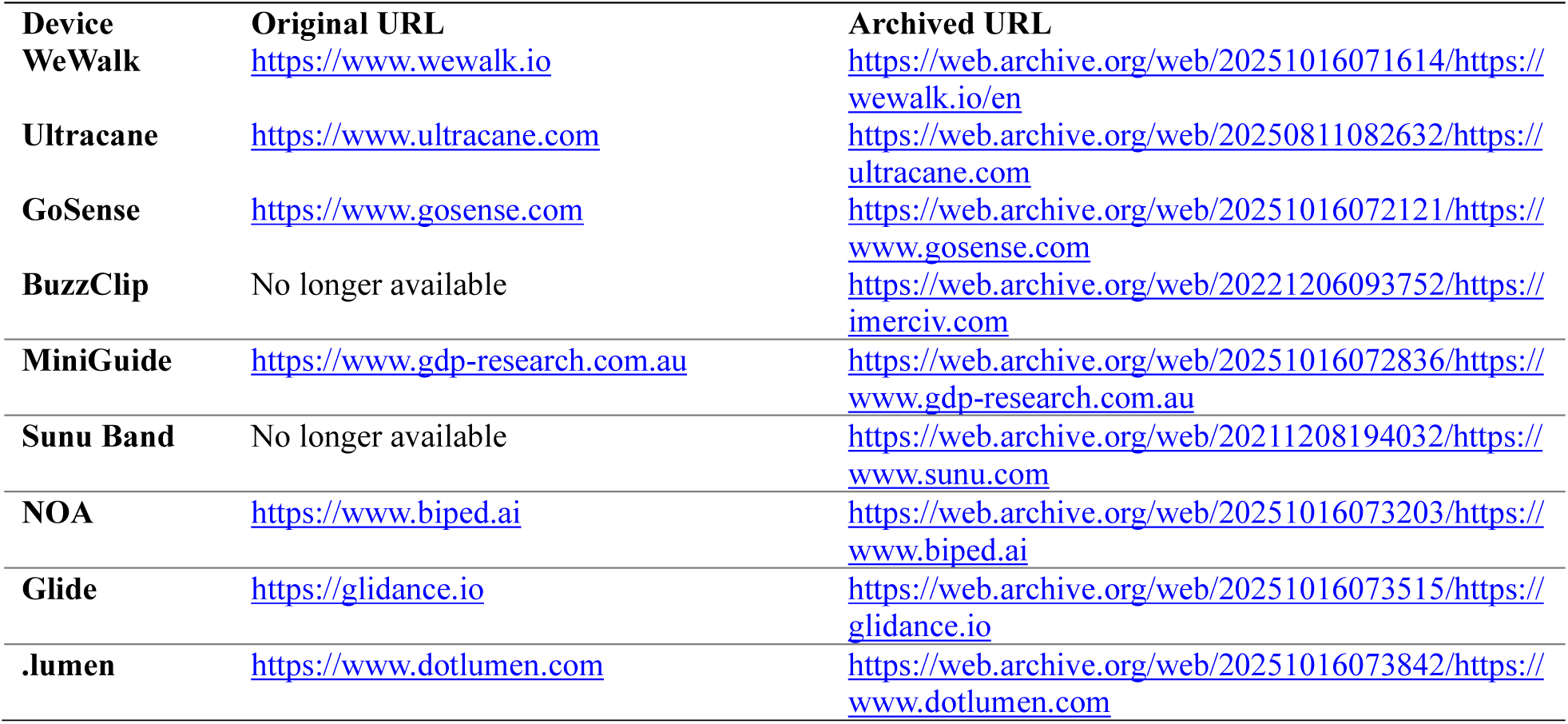
Overview of ETA devices with their original and archived Internet Archive URLs. Archived links ensure continued accessibility if original websites become unavailable. The BuzzClip and Sunu Band were discontinued at the time of the study.

| Device | Original URL | Archived URL |
| --- | --- | --- |
| WeWalk | https://www.wewalk.io | https://web.archive.org/web/20251016071614/https://wewalk.io/en |
| Ultracane | https://www.ultracane.com | https://web.archive.org/web/20250811082632/https://ultracane.com |
| GoSense | https://www.gosense.com | https://web.archive.org/web/20251016072121/https://www.gosense.com |
| BuzzClip | No longer available | https://web.archive.org/web/20221206093752/https://imerciv.com |
| MiniGuide | https://www.gdp-research.com.au | https://web.archive.org/web/20251016072836/https://www.gdp-research.com.au |
| Sunu Band | No longer available | https://web.archive.org/web/20211208194032/https://www.sunu.com |
| NOA | https://www.biped.ai | https://web.archive.org/web/20251016073203/https://www.biped.ai |
| Glide | https://glidance.io | https://web.archive.org/web/20251016073515/https://glidance.io |
| .lumen | https://www.dotlumen.com | https://web.archive.org/web/20251016073842/https://www.dotlumen.com |

**Table S2.**
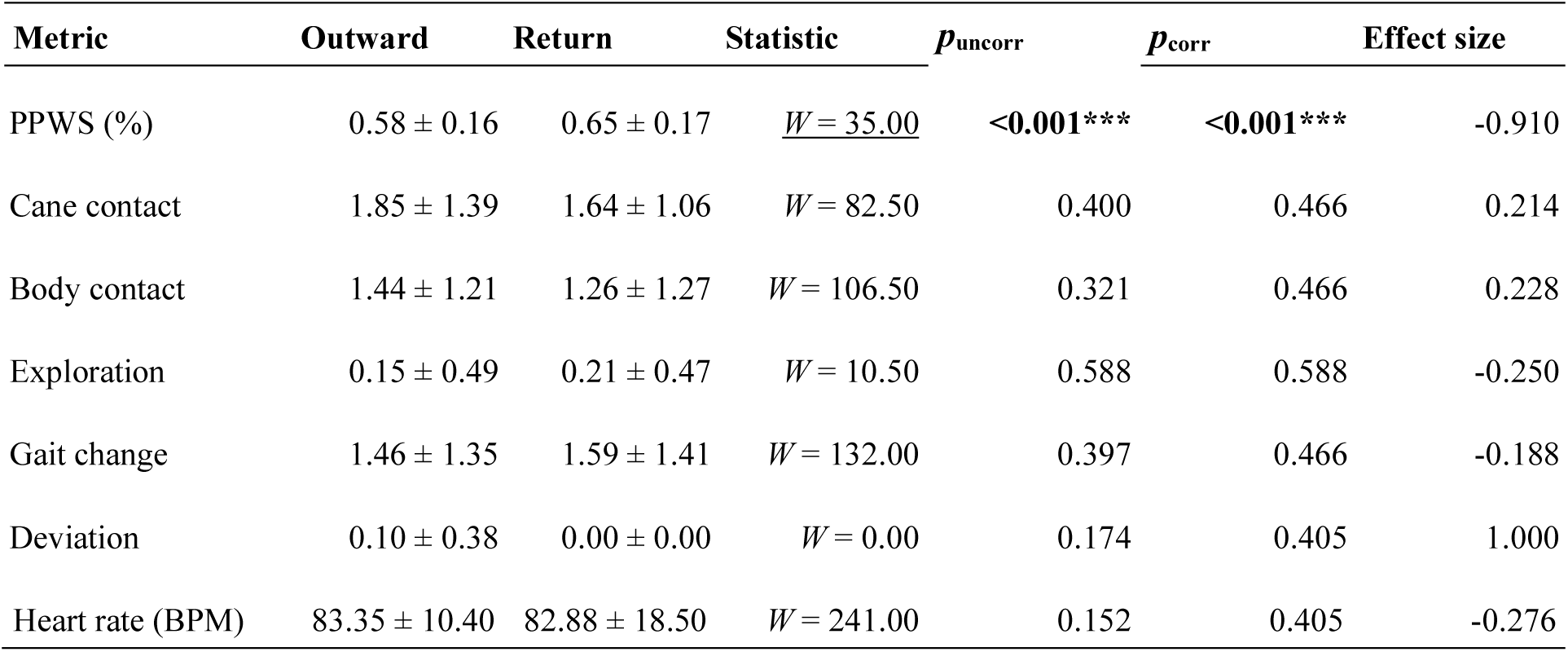
Performance metric differences between outward and return trials in the obstacle detection task. Wilcoxon signed-rank tests were performed as data was not normal. *W* values are reported. Values are presented as mean ± standard deviation (M ± SD). PPWS is expressed in percentage (%). All other values are event occurrences except heart rate, measured in beats per minute. Effect sizes are rank biserial correlation *rrb. P*-values are reported both uncorrected (*puncorr*) and corrected for multiple comparisons using the Benjamini–Hochberg procedure (*pcorr*).

| Metric | Outward | Return | Statistic | $p_{uncorr}$ | $p_{corr}$ | Effect size |
| --- | --- | --- | --- | --- | --- | --- |
| PPWS (%) | 0.58 ± 0.16 | 0.65 ± 0.17 | <u>$W = 35.00$</u> | **<0.001\*\*\*** | **<0.001\*\*\*** | -0.910 |
| Cane contact | 1.85 ± 1.39 | 1.64 ± 1.06 | $W = 82.50$ | 0.400 | 0.466 | 0.214 |
| Body contact | 1.44 ± 1.21 | 1.26 ± 1.27 | $W = 106.50$ | 0.321 | 0.466 | 0.228 |
| Exploration | 0.15 ± 0.49 | 0.21 ± 0.47 | $W = 10.50$ | 0.588 | 0.588 | -0.250 |
| Gait change | 1.46 ± 1.35 | 1.59 ± 1.41 | $W = 132.00$ | 0.397 | 0.466 | -0.188 |
| Deviation | 0.10 ± 0.38 | 0.00 ± 0.00 | $W = 0.00$ | 0.174 | 0.405 | 1.000 |
| Heart rate (BPM) | 83.35 ± 10.40 | 82.88 ± 18.50 | $W = 241.00$ | 0.152 | 0.405 | -0.276 |

**Table S3.** Responses to ternary-choice questions in the obstacle detection task. McNemar’s test was used to assess differences between conditions. Percentages reflect the proportion of participants who answered “yes” for each question under each condition. Δ% Yes indicates the difference in “yes” responses between NOA and BuzzClip. *P*-values are reported both uncorrected (*puncorr*) and corrected for multiple comparisons using the Benjamini–Hochberg procedure (*pcorr*).

| Question | %Yes NOA | %Yes BuzzClip | $\Delta\%$ Yes | $p_{uncorr}$ | $p_{corr}$ |
| --- | --- | --- | --- | --- | --- |
| Complement to cane | 100 | 53.85 | 46.15 | **0.031\*** | **0.047\*** |
| Increased mobility on familiar routes | 61.54 | 38.46 | 23.08 | 0.375 | 0.375 |
| Increased mobility on unfamiliar routes | 76.92 | 23.08 | 53.85 | **0.016\*** | **0.047\*** |

**Table S4.** Summary of participants’ responses to open-ended questions regarding learning difficulties, advantages, and disadvantages of NOA and the BuzzClip. Numbers in parentheses indicate the number of participants mentioning each aspect.

| Question | NOA | BuzzClip |
| --- | --- | --- |
| **Difficult learning aspects** | None (6), difficulty with sound spatialization (4), difficulty with device setting (3) | None (7), weak vibrations (4), imprecision (2) |
| **Mobility advantages** | Precision (8), easy interpretation (4), AI functionalities (4), flexibility / all-in-one nature (3), added safety and independence (3) | Lightweight / size (7), simplicity (3), potential utility for deaf-blind and children (1) |
| **Mobility disadvantages** | Weight / size (8), difficult interpretation of cues (4), none (2) | Weak vibrations (6), frequent vibrations (5), imprecision (5), none (1) |

**Table S5.** Data on participants demographics collected during the initial questionnaire. .

| Demographic | Summary |
| --- | --- |
| Age (years) | mean = 55.8 (SD = 19.8), range: 25 – 85 |
| Sex | 8 M (61.5%), 5 F (38.5%) |
| Vision level | 5 low vision (38.5%), 4 total (30.8%), 4 severe (30.8%) |
| Congenital | 9 no (69.2%), 4 yes (30.8%) |
| Employment status | 5 retired (38.5%), 4 employed (30.8%), 4 unemployed (30.8%) |
| Living situation | 8 family (61.5%), 5 alone (38.5%) |
| Living location | 8 urban (61.5%), 5 rural (38.5%) |
| Frequency on familiar routes | 7 daily (53.8%), 3 several times a week (23.1%), 2 weekly (15.4%), 1 rarely (7.7%) |
| Frequency on unfamiliar routes | 5 rarely (38.5%), 4 never (30.8%), 3 several times a week (23.1%), 1 weekly (7.7%) |

